# Comparative effects of exercise, nicotinamide riboside and time-restricted eating on aging-related biology in older adults: a randomized, controlled trial

**DOI:** 10.64898/2026.11.26.26369070

**Authors:** Magnus Berglind, Haithem Alkhefagie, Simon Reves, Jonas Bach Garbrecht, Brenna Osborne, Michael Petr, Aleksandr Dekan, Michael Ben Ezra, Indra Heckenbach, Garik Mkrtchyan, Kristoffer Tøttrup Norheim, Morten Scheibye-Knudsen

**Affiliations:** Center for Healthy Aging, Department of Cellular and Molecular Medicine, University of Copenhagen, Copenhagen, Denmark

**Author notes:** Correspondence: Morten Scheibye-Knudsen. Department of Clinical Research, Copenhagen University Hospital Amager & Hvidovre, 2650, Hvidovre, Denmark (Magnus Berglind). School of Biotechnology and Biomolecular Sciences, University of New South Wales, Kensington, NSW 2052, Australia. Senescope, Santa Cruz, CA 95060, USA.

## Abstract

Whether interventions targeting distinct processes of aging produces coordinated changes across human biology remains unknown. In this four-arm, open-label randomized trial (NCT05593939), 80 healthy adults aged ≥65 years were assigned to 12 weeks of aerobic exercise (EXE), nicotinamide riboside (NR), time-restricted eating (TRE) or control. 60 participants were analyzed per protocol. Each intervention produced distinct biological responses. NR markedly increased whole-blood NAD⁺, while TRE substantially reduced total and trunk fat without detectable muscle loss. Exercise, NR and TRE were associated with nominally lower age-acceleration estimates in selected DNA methylation-based aging measures, although responses varied across clocks. No significant intervention effects were detected for the primary outcome, plasma interleukin-6, or other circulating inflammatory markers. Integrative analyses revealed coordinated changes within biological domains and modest associations between transcriptional pathway activity and phenotypic changes, particularly among system-specific aging measures. Together, these findings suggest that distinct interventions can modify specific components of aging-related biology while revealing partially shared molecular responses, highlighting the multidimensional nature of human aging.

## Main

Geromedicine seeks to modify the shared biological processes underlying age-associated diseases, including chronic inflammation, metabolic dysregulation, and loss of cellular and tissue resilience^1,2^. Although interventions can modify aging phenotypes in animal models, evidence from randomized controlled trials in humans remains limited^3^. Aerobic exercise^4^, supplementation of the nicotinamide adenine dinucleotide (NAD^+^) precursor nicotinamide riboside (NR)^5^, and time-restricted eating (TRE)^6^ are biologically distinct interventions that engage different but interconnected aging processes. Human studies indicate that these interventions can improve overlapping metabolic and inflammatory outcomes^6–10^, while preclinical evidence suggests broader effects on aging-related pathways and healthspan phenotypes^11–15^. Together, they engage energy and NAD⁺ metabolism, nutrient sensing, inflammatory regulation and cellular maintenance and repair: processes that deteriorate with age^16^, and contribute to multimorbidity, frailty, and other age-related conditions^17–19^. We hypothesize that these interventions may impact similar aging processes; however, the specificity and magnitude of the effects remain unresolved.

To test this, we conducted a four-arm, 12-week randomized controlled trial in 80 healthy adults aged ≥65 years, randomly assigned to aerobic exercise, NR (2 g daily), TRE or control. Because chronic low-grade inflammation is a hallmark of aging and a driver of chronic diseases^20,21^, plasma interleukin-6 (IL-6) was prespecified as the primary outcome. Prespecified secondary and exploratory outcomes spanning biochemical, physiological, functional, biological-aging and transcriptomic domains addressed the broader geroscience question. We found that each intervention elicited distinct biological responses, including changes in NAD⁺ metabolism, body composition and selected biological-aging measures. Interestingly, despite differences in physiological responses, transcriptomic analyses revealed partially overlapping molecular adaptations. Together, these findings provide insight into the specificity and convergence of human responses to interventions targeting aging and highlight the importance of evaluating aging biology across multiple biological domains.

## Results

### Participant recruitment, baseline characteristics, and safety

Of 138 individuals screened, 80 (40 women; mean age, 71.0 years) were randomized in sex-stratified blocks to CTRL, EXE, NR or TRE (n = 20 per group; Fig. 1a). Four participants did not complete follow-up (NR, n = 1; TRE, n = 3). The per-protocol population comprised 60 participants (CTRL, n = 11; EXE, n = 18; NR, n = 17; TRE, n = 14), with exclusions detailed in Fig. 1a. Notably, six controls were excluded due to substantial fat loss (>10%), inconsistent with maintenance of habitual lifestyle, and thus excluded due to failure to adhere to study regimen. Baseline characteristics were similar across groups (per-protocol; Table 1). The trial design and assessment schedule are shown in Fig. 1b. Adverse events were reported by 5 participants (25%) in CTRL, 6 (30%) in EXE, 15 (75%) in NR and 10 (50%) in TRE (Supplementary Table 1).

**Fig. 1.**
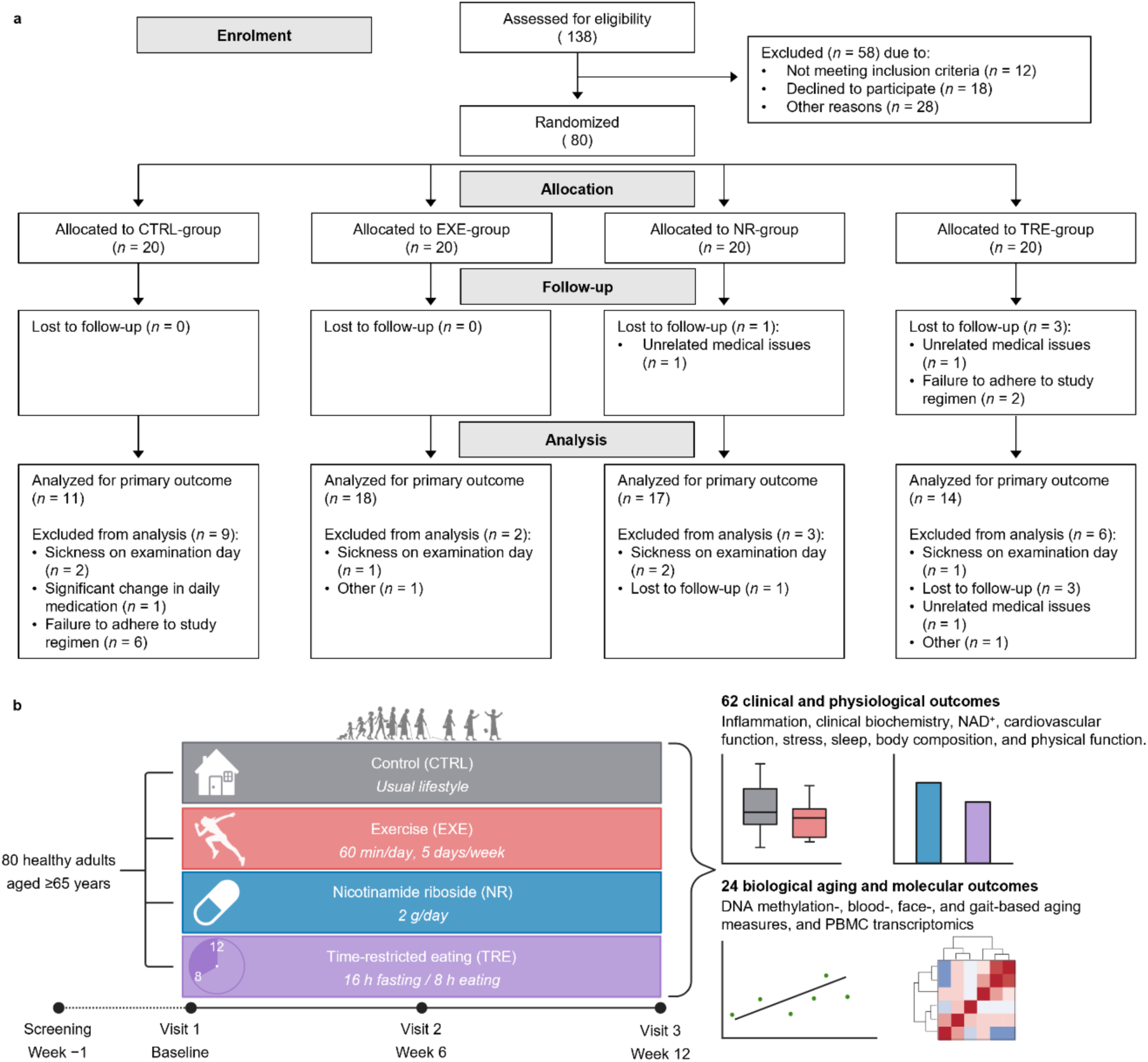
CONSORT flow diagram and study design. **a,** CONSORT flow diagram for the four-arm randomized controlled trial in healthy older adults (aged ≥65 years). Eighty participants were randomized (*n* = 20 per group) to control (CTRL), aerobic exercise (EXE), nicotinamide riboside (NR) or time-restricted eating (TRE). The primary analyses were performed in the per-protocol analysis set. Exclusions due to loss to follow-up and protocol deviations are shown. **b**, Study design schematic showing intervention arms, visit schedule and outcome domains.

**Table 1.** Participant characterization.

| **Characteristic** | **CTRL** | **EXE** | **NR** | **TRE** |
|---|---|---|---|---|
|  | (n = 11) | (n = 18) | (n = 17) | (n = 14) |
| **Age, years** | 68.4 (67.4-71.4) | 70.0 (67.5-75.5) | 71.0 (68.1-75.6) | 70.6 (68.7-72.2) |
| **Female sex, n** | 5 (45%) | 10 (56%) | 8 (47%) | 6 (43%) |
| **Weight, kg** | 79.8 (74.5-89.9) | 76.5 (66.4-85.7) | 74.4 (70.4-84.0) | 79.7 (68.1-86.9) |
| **Height, cm** | 173.0 (164.0-177.0) | 171.0 (165.0-178.0) | 168.0 (162.0-176.0) | 170.0 (160.0-178.0) |
| **BMI, kg/m<sup>2</sup>** | 27.1 (24.3-31.5) | 25.4 (23.2-28.9) | 26.5 (24.6-29.9) | 26.5 (24.9-28.2) |
| **Fat mass, kg** | 22.9 (15.7-32.4) | 21.3 (16.7-27.1) | 20.5 (17.6-26.2) | 19.8 (15.7-24.8) |
| **Muscle mass, kg** | 56.7 (49.0-60.9) | 51.3 (44.6-56.9) | 52.8 (43.2-57.8) | 53.7 (42.0-61.8) |

Per protocol population (*n* = 60). Continuous: median (Q1–Q3); Categorical: n (%).

### Interventions produce clear but distinct biochemical and physiological responses

We first examined NAD⁺ levels, body composition and physical activity as measures of the anticipated responses to NR, TRE and EXE, respectively. NR produced clear biochemical target engagement: after adjustment for baseline, post-intervention protein-normalized whole-blood NAD⁺ was 34.69 pmol mg ⁻¹ higher following NR than CTRL (95% CI, 26.40 to 42.97; P < 0.001; Fig. 2a and Supplementary Table 2). Within the NR group, NAD⁺ increased by 36.37 ± 18.86 pmol mg⁻¹ (P < 0.001), whereas it remained stable in CTRL, EXE and TRE. In exploratory cross-sectional analyses at baseline, whole-blood NAD⁺ was not associated with chronological age, consistent with recent reports questioning a uniform age-related decline in circulating NAD⁺^22–24^ (Supplementary Fig. 1a and Supplementary Table 3). Complete analyses for all outcomes are provided in Supplementary Table 2.

**Fig. 2.**
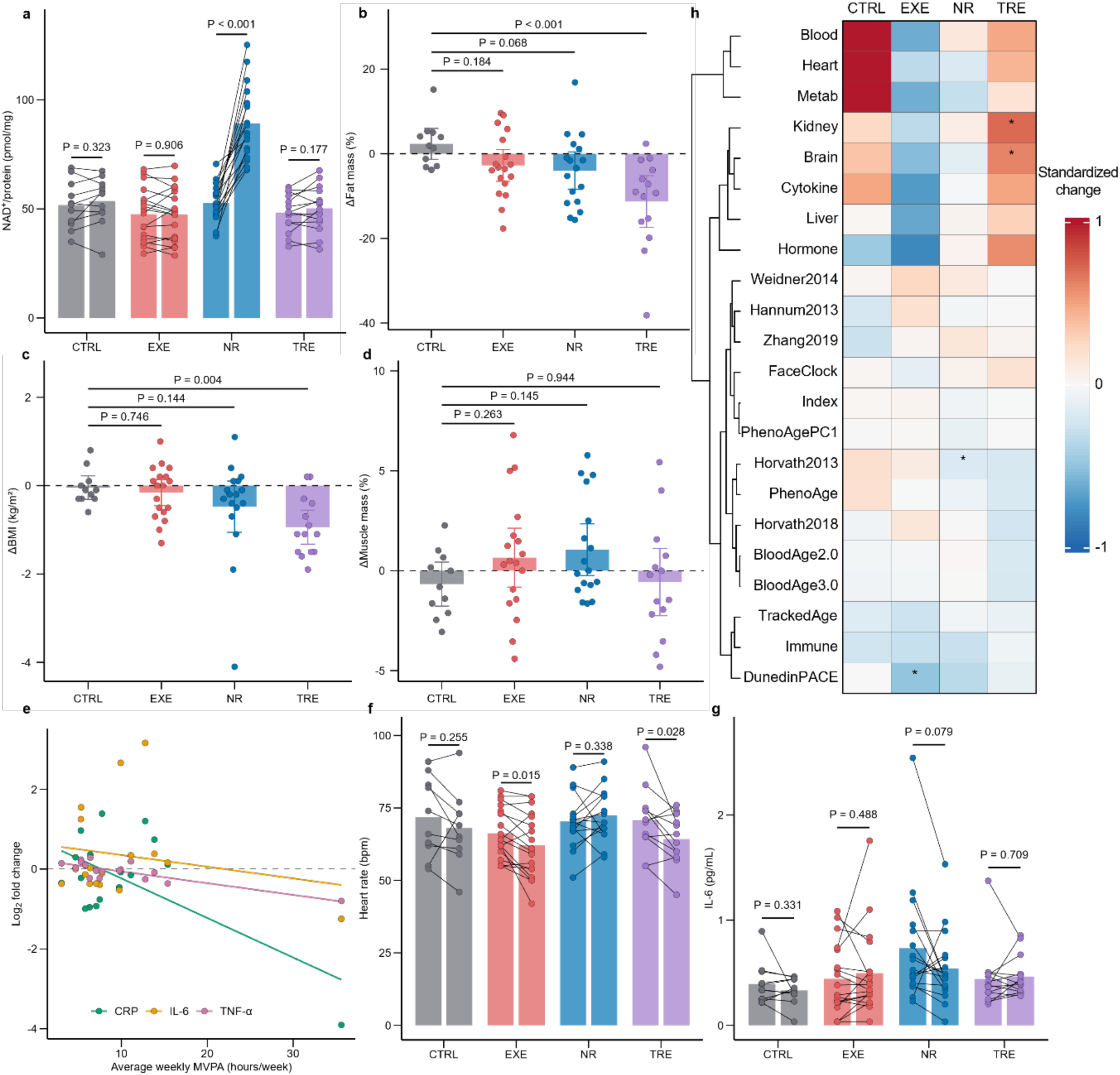
Effects of the interventions on inflammation, NAD⁺, biological aging and physiological outcomes. **a**, Paired pre- and post-intervention whole-blood NAD⁺. **b**–**d**, Changes in fat mass (**b**), BMI (**c**) and BIA-estimated muscle mass (**d**), shown as percentage changes from baseline for fat and muscle mass and absolute change for BMI. **e**, MVPA vs pre-to post-intervention log_2_ fold change in CRP, IL-6 and TNF-α among participants in the EXE group with available Fitbit data. Points represent individual participants and solid lines show marker-specific linear regression fits for visualization. The horizontal dashed line indicates no change from baseline. Weekly MVPA was estimated from Fitbit recordings during the final 31 days of the intervention. **f**, Paired pre- and post-intervention resting heart rate. **g**, Paired pre- and post-intervention IL-6. **h**, Heatmap of standardized pre-to post-intervention changes in biological-aging measures across intervention arms, expressed in units of the baseline standard deviation. Red indicates an increase and blue decrease in the respective aging measure; asterisks denote unadjusted two-sided Wilcoxon signed-rank P < 0.05. Bars in **a**, **f** and **g** show group means at each time point, whereas bars in **b**–**d** show mean change from baseline. Points represent individual participants, paired lines connect repeated measurements where applicable, and error bars show 95% confidence intervals. *P* values in **a**, **f** and **g** indicate within-group paired comparisons. Between-group P values in **b** and **d** were derived from ANCOVA models of log-transformed post-intervention values adjusted for log-transformed baseline values and sex; P values in **c** were derived from ANCOVA of post-intervention BMI adjusted for baseline BMI and sex.

Among the prespecified physiological outcomes, TRE produced a distinct response in body composition (Fig. 2b). Total fat mass decreased by 11.27 ± 10.63% in TRE but increased by 2.34 ± 5.44% in CTRL (adjusted difference, −13.76%; 95% CI, −19.62 to −7.48; P < 0.001; Supplementary Table 2). In TRE, BMI decreased by 0.94 ± 0.66 kg m⁻² (adjusted difference versus CTRL, −0.92 kg m⁻²; 95% CI, −1.54 to −0.30; P = 0.004; Fig. 2c), and trunk fat mass decreased by 13.61 ± 14.26% (adjusted difference, −16.41%; 95% CI, −24.29 to −7.70; P < 0.001; Supplementary Table 2) without detectable loss of bioelectrical impedance analysis (BIA)-estimated muscle mass (adjusted difference, −0.07%; 95% CI, −2.16 to 2.05; P = 0.944; Fig. 2d). Neither EXE nor NR showed statistically detectable differences in fat mass, muscle mass or BMI relative to CTRL. In sensitivity analyses including all participants with available clinical data, the TRE-associated reduction in total fat mass persisted, although attenuated (adjusted difference, −8.15%; 95% CI, −14.00 to −1.90; P = 0.012), whereas the trunk fat-mass difference was smaller and no longer statistically detectable (−7.91%; 95% CI, −16.99 to 2.17; P = 0.118; Supplementary Table 4). Objective monitoring confirmed substantial exercise exposure in EXE: among 17 participants with Fitbit data from the final 31 days, median moderate-to-vigorous physical activity (MVPA) was 448 min per week (IQR, 347–666), with 15 (88.2%) averaging ≥300 min per week and ≥60 min of MVPA recorded on a median of 4.1 days per week (Supplementary Table 5).

Resting heart rate decreased within EXE and TRE, consistent with established effects of aerobic exercise^25^ and recent evidence suggesting modest reductions with TRE^26^ (Fig. 2f and Supplementary Table 2). Exploratory analyses within NR further suggested that greater increases in NAD⁺ were associated with greater reductions in resting heart rate (ρ = −0.51, P = 0.050; Supplementary Fig. 1b). Thus, the interventions were active, but their most prominent short-term responses emerged in different biological domains.

### Inflammatory responses to exercise, NR and TRE

Previous studies have reported anti-inflammatory effects of NR^7,8^, fasting^10^ and exercise^4^. Surprisingly, the prespecified primary outcome IL-6 did not differ between any intervention group and CTRL after adjustment for baseline IL-6 at 12 weeks (Fig. 2g and Supplementary Table 2). Although NR showed the largest within-group reduction (−0.19 ± 0.42 pg ml⁻¹) consistent in direction with other literature on NAD^+^ precursor supplementation^8,27^, the adjusted difference versus CTRL was 0.05 pg ml^−1^ (95% CI, −0.16 to 0.25; P = 0.643). EXE and TRE likewise did not differ from CTRL. The prespecified secondary inflammatory outcomes CRP and TNF-α also showed no detectable differences between the intervention groups and CTRL (Supplementary Table 2). In exploratory within-EXE analyses, greater weekly MVPA was significantly associated with larger reductions in TNF-α, surviving FDR correction (Fig. 2e and Supplementary Table 6). Together, these findings indicate that none of the interventions produced a consistent systemic anti-inflammatory response over 12 weeks, although the EXE findings suggest that inflammatory responses may vary with achieved exercise exposure.

### Biological aging measures show heterogeneous intervention-response patterns

To test whether the interventions converged on downstream biological aging measures, we assessed DNAm-derived aging measures in PBMCs, alongside exploratory biological age measures derived from blood biochemistry^28^, facial photographs^29^, and gait analysis (TrackedAge™). At baseline, chronological age was positively associated with several DNAm-derived aging measures, particularly the Metabolic, Immune and Brain system-specific clocks, as well as established epigenetic clocks and the facial aging measures FaceClock and FaceRisk (Supplementary Fig. 1c and Supplementary Table 3). Among conventional outcomes, older age was associated with higher immature granulocyte counts, lower total and LDL cholesterol concentrations, and reduced handgrip strength. These associations remained significant after adjustment for sex and correction for multiple testing (FDR q < 0.05). In contrast, systemic inflammatory markers, NAD⁺ concentrations, body composition and gait-related outcomes showed no significant associations with chronological age after FDR correction (Supplementary Fig. 1c and Supplementary Table 3). Within-group changes varied across measures and intervention arms (Fig. 2h). In ANCOVA models adjusted for baseline and sex (Supplementary Table 2), post-intervention age accelerations, expressed in years, were lower following EXE than CTRL for the Metabolic (−4.15; 95% CI, −6.78 to −1.52; P = 0.003), Blood (−16.97; 95% CI, −29.37 to −4.58; P = 0.008), Cytokine (−6.06; 95% CI, −10.49 to −1.63; P = 0.008), Brain (−4.58; 95% CI, −8.93 to −0.24; P = 0.039) and Heart clocks (−11.63; 95% CI, −22.83 to −0.44; P = 0.042). However, this pattern was not uniform across measures: Hannum2013 was higher in EXE than CTRL (2.14; 95% CI, 0.20 to 4.08; P = 0.032), while Zhang2019 showed a similar direction (1.16; 95% CI, −0.09 to 2.41; P = 0.069). NR showed a partially overlapping pattern, with lower Metabolic (−3.58; 95% CI, −6.20 to −0.95; P = 0.009) and Blood clock estimates (−13.48; 95% CI, −25.81 to −1.14; P = 0.033), but a higher Zhang2019 estimate (1.35; 95% CI, 0.09 to 2.60; P = 0.036), relative to CTRL. TRE showed lower Metabolic (−2.90; 95% CI, −5.73 to −0.06; P = 0.045) estimates as well as lower estimates across several first-generation clocks, most pronounced for Horvath2013 (−2.59; 95% CI, −5.03 to −0.15; P = 0.038). Sensitivity analyses adjusting for changes in estimated PBMC cell composition yielded largely consistent, suggesting that these data were not explained by redistribution of immune-cell populations (Supplementary Table 7). Overall, the interventions produced heterogeneous, measure-dependent signals rather than a uniform shift across biological-aging measures. Thus, biochemical and physiological responses did not translate into a common short-term aging-biomarker signature.

### Changes in aging-related outcomes cluster within biological domains

To examine whether changes across outcomes were coordinated, we performed an exploratory analysis of Spearman correlations among pre-to-post changes in 85 physiological, biochemical, functional and biological-aging measures, pooling participants across intervention groups. Measures were grouped into related outcome domains, such as DNAm aging, clinical biochemistry, and inflammation and hematology, as illustrated in Fig. 3a. Of 3,570 pairs tested, 104 met the thresholds of |ρ| ≥ 0.40 and FDR-adjusted P ≤ 0.10 (pairwise *n* = 55–60), of which 85 were within-domain and 19 cross-domain (Fig. 3a, Supplementary Table 8). DNAm-based measures formed the dominant network: among nine system-specific clocks, 30 of 36 possible correlations were retained (ρ = 0.43–0.84). Cross-domain associations were less frequent and included links between changes in urea, BloodAge2 and gait measures, whereas CRP, IL-6 and TNF-α were absent from the retained network. Baseline outcome correlations are shown in Supplementary Fig. 2, while correlations between pre-to post-intervention changes are shown in Supplementary Fig. 3. In this pooled analysis, short-term changes were coordinated predominantly among measures within the same outcome domain, with comparatively fewer cross-domain associations. This was consistent with the intervention-level findings above, in which responses in individual domains were not accompanied by broad coordinated changes across others.

**Figure 3.**
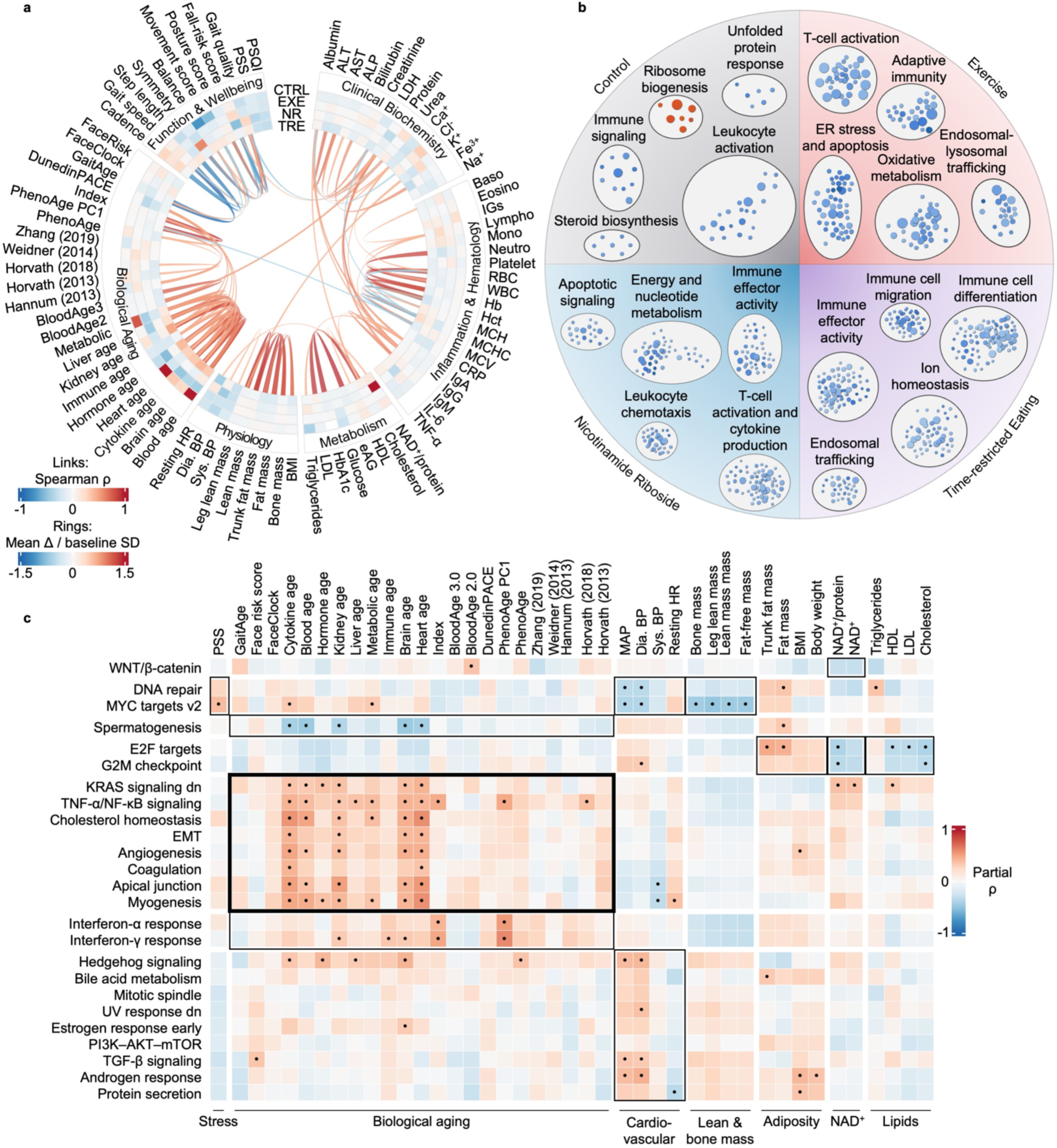
Relationships between study outcomes and transcriptomic responses to the interventions. **a,** Spearman correlations between pre-to post-intervention changes across study outcomes, pooled across groups. Sectors represent outcomes grouped by domain; links indicate correlations with |ρ| ≥ 0.40 and FDR-adjusted P ≤ 0.10, with color indicating direction and width indicating magnitude. Concentric rings show mean changes for CTRL, EXE, NR and TRE, standardized to the baseline SD. **b,** EnrichmentMap networks of within-group transcriptomic changes, showing the five largest pathway clusters per intervention (GSEA FDR ≤ 0.25). Nodes represent GO Biological Process gene sets, colored by normalized enrichment score (blue, negative; orange, positive); ellipses indicate pathway clusters. **c,** Associations between pre-to post-intervention changes in Hallmark pathway activity (ssGSEA) and phenotypic outcomes among 45 participants with paired RNA-sequencing data. Colors indicate partial Spearman correlations adjusted for intervention, age, sex and baseline outcome values (blue, negative; red, positive); dots indicate P < 0.05. Gene sets (rows) are grouped into 10 clusters based on their co-variation across participants, and outcomes (columns) into 15 measurement domains. Associations between pathways and outcomes were evaluated using permutation tests of mean squared partial correlations (20,000 permutations within interventions), with FDR correction across 150 combinations. Thin outlines indicate P < 0.05; thick outline indicates meeting FDR threshold of q < 0.25 (P = 0.0015, q = 0.23). For clarity, 25 of 50 gene sets and 42 of 97 outcomes are displayed. Complete data can be found in Supplementary Table 9.

### Transcriptomic changes suggest partial molecular convergence

To examine whether molecular responses showed a different convergence than the higher-order phenotypic responses, we performed gene set enrichment analysis (GSEA) of within-arm pre-to-post changes in bulk RNA-sequencing of PBMCs and summarized enriched Gene Ontology Biological Process terms using EnrichmentMap networks (FDR < 0.25; Fig. 3b). Immune-related processes recurred across EXE, NR and TRE, whereas cellular-stress and apoptosis programs were prominent in EXE and NR. Metabolic processes were represented in both EXE and NR, while endolysosomal or endosomal trafficking was represented in both EXE and TRE. CTRL also showed predominantly negative enrichment of immune- and stress-related pathways, alongside positive enrichment of ribosome-biogenesis pathways. Thus, although these within-arm analyses do not establish randomized intervention effects, the recurrence of immune-related pathways across EXE, NR and TRE suggests that immune regulation may be a common point of molecular convergence, while other pathway responses remained more intervention-specific.

### Associations between gene expression and aging-related outcomes

We next examined whether changes in PBMC transcriptional pathways were associated with concurrent changes in clinical, functional and biological-aging outcomes. Among 45 participants with paired RNA-sequencing data, we assessed associations between changes in 50 Hallmark gene sets and 97 phenotypic outcomes, adjusting for intervention group, age, sex and baseline outcome values (Fig. 3c). Across 4,850 pathway–outcome pairs, overall coupling was modestly greater than expected by chance (mean squared partial correlation, 0.032 versus 0.027 under permutation; P = 0.031). However, no individual association survived FDR correction (all q ≥ 0.28; Supplementary Table 9), suggesting that the overall relationship was distributed across numerous modest associations rather than driven by a few individually robust correlations.

To examine these relationships at a broader biological level, we grouped transcriptional pathways into ten clusters based on their co-variation across participants, and phenotypic outcomes into 15 measurement domains. Of 150 pathway-cluster x outcome-domain combinations, 13 showed significant associations, and one met an exploratory FDR threshold of q < 0.25 (Fig. 3c). This association linked biological-aging measures to a cluster containing TNF-α signaling via NF-κB, cholesterol homeostasis and other signaling pathways (P = 0.0015, q = 0.23). Notably, the relationship was strongest for six system-specific DNAm aging clocks (Heart, Cytokine, Brain, Kidney, Blood and Metabolic), whereas conventional epigenetic clocks showed weaker associations. The relationship was directionally consistent across intervention arms, although overall RNA–phenotype coupling did not differ significantly between arms (P = 0.14).

Together, these findings indicate that changes in gene-expression pathways were associated with changes in aging-related outcomes more than expected by chance, particularly among system-specific DNAm clocks. Alongside the predominantly within-domain phenotypic correlations (Fig. 3a) and recurring immune-related transcriptional responses (Fig. 3b), these findings raise the possibility of partially overlapping molecular processes despite distinct physiological and aging-related responses to the interventions.

## Discussion

In this randomized trial of healthy older adults, exercise, NR and TRE produced distinct responses, but no common short-term signature emerged across aging-related outcomes. EXE participants accumulated substantial MVPA, NR markedly increased whole-blood NAD⁺, and TRE reduced total and trunk fat without detectable BIA-estimated muscle loss. Yet no intervention produced statistically detectable differences in IL-6, CRP or TNF-α relative to CTRL, biological-aging measures moved in heterogeneous directions, and changes across the wider phenotype were coordinated mainly within biological domains. This is notable because the interventions act on processes that are thought to interact within aging biology. Our findings suggest that such interconnection does not mean that affecting one process will readily drive corresponding change in others. Instead, the findings are consistent with a hierarchy of response: interventions may first alter biology closest to their point of action, whereas broader effects may require sufficient duration, access to the relevant tissues, and correction of a biological deficit that itself limits physiology.

NR and TRE illustrate this distinction. NR robustly increased whole-blood NAD⁺, consistent with previous human supplementation studies^7,8,27,30,31^, but this did not translate into lower circulating inflammatory markers relative to CTRL. Increasing the pool of circulating NAD⁺ does not imply equivalent repletion across tissues or that NAD⁺ availability was limiting downstream inflammatory processes in these participants. This is consistent with growing evidence that age-related NAD⁺ decline is context-dependent^24,32,33^ rather than a universal feature of aging^22–24^. Conversely, TRE produced a distinct physiological response, with marked adiposity loss without detectable muscle loss, yet no consistent improvement across inflammatory or biological-aging measures. The DNAm clock analyses further highlighted this heterogeneity: exercise mostly affected organ specific clocks while TRE impacts first generation clocks to a higher degree. These findings suggest that interconnected biological systems can accommodate substantial local change without necessarily undergoing coordinated remodeling elsewhere.

The integrative analyses revealed further differences in how biological changes were related. Phenotypic changes were predominantly correlated within domains, particularly among DNAm aging measures, with relatively few associations across different physiological systems. In contrast, immune- and stress-related transcriptional pathways recurred across intervention groups, and changes in PBMC gene expression were associated with phenotypic changes more than expected by chance. The most prominent relationship involved inflammatory and metabolic gene-expression pathways and several system-specific DNAm clocks, whereas associations with conventional epigenetic clocks were weaker. This raises the possibility that different DNAm aging measures capture distinct aspects of short-term molecular changes. However, both RNA expression and DNAm were measured in PBMCs, and these associations do not necessarily indicate corresponding changes in the organs represented by the system-specific clocks. This suggests that molecular changes may be related even when broader physiological responses are different. The recurring transcriptional pathways across interventions are consistent with some shared molecular responses, although similar changes were also observed in CTRL. Moreover, RNA–phenotype associations did not differ detectably between intervention groups. Whether these molecular relationships eventually translate into broader physiological changes, or primarily reflect changes within circulating immune cells, remains to be established.

The study has several limitations. The modest and uneven per-protocol groups limited precision, particularly for individual-arm comparisons, and post-randomization exclusions may have introduced selection bias. Sensitivity analyses using all available clinical data addressed this concern, although molecular analyses remained restricted to the per-protocol cohort. The open-label design, including the absence of an NR placebo, may have influenced participant behavior, while hydration-sensitive BIA measures may have affected body composition estimates. Multiple exploratory comparisons increased the risk of chance findings. Furthermore, 12 weeks may have been insufficient to capture slower biological responses, and blood-based measures may miss tissue-specific effects, particularly in relatively healthy participants with fewer correctable deficits.

Despite these constraints, our comparative design demonstrates that short-term geroscience interventions can substantially modify specific components of human aging-related biology. The associations between transcriptional pathways and system-specific aging measures further suggest that molecular responses may reveal shared biological processes not readily captured by conventional physiological outcomes. These findings highlight the value of integrating complementary molecular and physiological measures to characterize intervention responses and provide a framework for identifying which aspects of aging biology are amenable to modification in humans.

## Methods

### Ethical approval, informed consent, and study location

The study was approved by the regional ethical review board for the capital region of Denmark (H-21017723) and registered at ClinicalTrials.gov (NCT05593939). All procedures were conducted in accordance with the Declaration of Helsinki and the ethical principles of Good Clinical Practice. Written informed consent was obtained from all participants prior to enrolment. Clinical assessments were conducted between 17 May and 21 December 2022 at the University of Copenhagen and Rigshospitalet, Copenhagen, Denmark. The study protocol specified circumstances warranting early suspension or termination, including unacceptable participant risk, evidence of efficacy or futility, inadequate protocol compliance or data quality, or attainment of the primary endpoint. The trial followed CONSORT guidelines. The study protocol, including the statistical analysis plan, is provided as Supplementary File 1.

### Study design and participants

The SlowAge study was a four-arm, parallel-group, open-label randomized controlled trial. The trial was designed to evaluate the superiority of exercise, nicotinamide riboside supplementation and time-restricted eating over a no-intervention control in modifying aging-related outcomes in older adults. Eligible participants were aged ≥65 years, in good general health, willing and available to complete all study procedures for the duration of the trial and provided written informed consent. Participants were excluded if they were unable or unwilling to take oral supplements, adhere to the time-restricted eating regimen or perform the prescribed exercise intervention; were current smokers or had used nicotine products within the previous 10 years; chronically used supplements containing vitamin B or nicotinamide riboside; had received another investigational drug or intervention within the previous year; or had been diagnosed with cancer within the previous 5 years. A total of 80 participants (40 women and 40 men) were enrolled and randomized equally to control (CTRL), aerobic exercise (EXE), nicotinamide riboside supplementation (NR) or time-restricted eating (TRE). Participants followed their assigned intervention for 12 weeks, with study visits at baseline, week 6 and week 12. Concomitant medical care was not systematically recorded. No patients or members of the public were involved as research partners in the design, conduct or reporting of the trial.

### Randomization and analysis population

Following screening, participants were randomized 1:1:1:1 within sex strata using permuted blocks of 4 generated in Microsoft Excel. Each block was randomized only after all its participants had enrolled, ensuring that assignments were unknown during enrollment. Owing to the nature of the interventions, participants and study personnel were not blinded to group allocation. Outcome assessors, laboratory personnel and statistical analysts were also not blinded to allocation. The per-protocol analysis set comprised 60 participants (CTRL, n = 11; EXE, n = 18; NR, n = 17; TRE, n = 14). Participants were excluded from the per-protocol analysis because of loss to follow-up, protocol deviations, changes in medication or health status, or non-adherence to their study intervention, as detailed in Fig. 1a. DNA methylation and transcriptomic profiling were restricted to the per-protocol population, although clinical measurements remained available for excluded participants where collected. Primary analyses used the per-protocol population, with smaller sample sizes where specific measurements were unavailable. Sensitivity analyses of clinical outcomes included all randomized participants with available baseline and follow-up measurements, irrespective of protocol adherence.

### Intervention and adherence monitoring

All interventions were self-administered by participants, with adherence monitored remotely by study personnel. No specific intervention-provider or delivery-site eligibility criteria applied.

Control: Participants were instructed to maintain their habitual lifestyle and dietary patterns throughout the 12-week study period.

Exercise: Participants engaged in moderate-to-vigorous aerobic exercise five days per week. Exercise duration increased progressively from 30 min per day (week 1) to 45 min per day (week 2), reaching 60 min per day from week 3 onward. Heart rate targets were 60–90% of maximum heart rate, monitored continuously using a Fitbit device.

Nicotinamide riboside: Participants consumed 2 g NR per day (1 g in the morning and 1 g in the evening) in capsule form, taken with meals. Supplements were provided by Elysium Health (New York, NY, USA).

Time-restricted eating: Participants practiced a 16:8 fasting-to-eating ratio, abstaining from caloric intake for 16 h per day and consuming food ad libitum within an 8-hour eating window. The default schedule was 12:00–20:00 h, but participants could adjust timing to fit personal routines.

Participants were contacted weekly by telephone throughout the 12-week intervention to assess adherence, adverse effects and changes in lifestyle. Adverse effects were assessed through participant self-report during these contacts. Physical activity in EXE participants was monitored throughout the intervention by remote assessment of Fitbit data to support adherence to the prescribed exercise program. Quantitative adherence was assessed from available Fitbit recordings during the final 31 days of the intervention. Fitbit-derived MVPA data were available for 17 participants. Participants recorded a median daily MVPA of 62 min/day (IQR 49–92) and a median average weekly MVPA of 448 min/week (IQR 347–666); 15 of 17 participants (88.2%) averaged at least 300 min/week. Participants recorded ≥60 min MVPA on an estimated median of 4.1 days/week. Detailed adherence data are provided in Supplementary Table 5.

### Questionnaires

Participants completed standardized questionnaires assessing health status, diet, alcohol consumption, exercise habits, stress and sleep quality. Self-reported medical history at baseline is summarized in Supplementary Table 10, while dietary habits, alcohol consumption and physical activity at baseline, week 6 and week 12 are summarized in Supplementary Table 11.

### Anthropometric and physical function assessments

Height was measured once at baseline using a Soehnle Professional 5003 height rod (Soehnle, Germany). Body composition was assessed with bioelectrical impedance analysis using a MC 980MA-N Plus (Tanita, Denmark) while participants wore light clothing. Blood pressure and resting heart rate were measured in a seated position after a 10-minute rest using an ADC Advantage Ultra 6023N monitor (ADC, USA). Seated handgrip strength was evaluated as the mean of three maximal efforts from each hand in a seated position using a Baseline LITE hydraulic dynamometer (Fabrication Enterprises Inc., USA). Participants were video recorded during a 4-meter walk using off-the-shelf cameras. Videos were analyzed using the TrackedGait™ platform (Tracked Biotechnologies LLC, Manassas, Virginia, USA). The platform derived gait speed, cadence, step length, symmetry, balance, posture, movement, gait-quality and fall-risk scores. Measures other than gait speed, cadence, step length, and symmetry are reported as exploratory research measures. The participants were photographed using a Nikon D3500 high-resolution digital camera.

### Blood collection

Venous blood samples were collected after a 12-hour overnight fast at baseline (week 0) and follow-up (week 12) at the Department of Clinical Biochemistry, Rigshospitalet, Copenhagen, Denmark. A subset of whole-blood aliquots was transferred to the research laboratory for downstream analyses, while the remaining samples were retained at the hospital for clinical biochemistry and inflammatory marker assessment. Routine clinical biochemistry was analysed using automated cobas systems (Roche Diagnostics). IL-6 and TNF-α were quantified in EDTA plasma using electrochemiluminescence-based immunoassays (Meso Scale Diagnostics).

### PBMC isolation

Peripheral blood mononuclear cells (PBMCs) were isolated using Ficoll-Paque PLUS (Sigma-Aldrich, 17144002). The Ficoll reagent was gently inverted to ensure homogeneity, and 3 mL was added to 15 mL conical tubes. Whole blood (5 mL) diluted 1:1 with PBS was carefully layered on top of the Ficoll solution. Samples were centrifuged at 400 × g for 30 min at room temperature with low acceleration and no brake. The PBMC layer at the plasma–Ficoll interface was collected and washed in 10 mL PBS by centrifugation at 350 × g for 10 min at room temperature. The supernatant was discarded, and the pellet was gently resuspended in 4 mL PBS and divided into safe lock microtubes. Cells were pelleted again by centrifugation at 300 × g for 10 min at 4 °C, and the supernatant was removed. PBMC pellets were snap-frozen in liquid nitrogen and stored at −80 °C until further analysis.

### DNA methylation profiling and preprocessing

DNA extraction, bisulfite conversion and DNA methylation profiling were performed by Elysium Health (New York, NY, USA). Genomic DNA was isolated from peripheral blood mononuclear cell pellets collected at baseline and after the 12-week intervention and quantified using standard fluorometric methods. Following bisulfite conversion, genome-wide DNA methylation was measured using the Illumina Infinium MethylationEPIC BeadChip array. Raw signal intensities were processed by Elysium Health using a standardized pipeline that included background correction, normalization and probe-level quality control. Processed methylation beta values and biological-aging estimates derived from multiple DNA methylation-based algorithms were provided for downstream analysis. Subsequent quality assessment, derivation of age-acceleration measures and statistical analyses were performed independently at the University of Copenhagen.

### Biological age and age-acceleration measures

DNA methylation-derived biological aging measures included established epigenetic clocks developed by Horvath2013^34^, Horvath2018^35^, Hannum2013^36^, Weidner2014^37^, Zhang2019^38^, DNAmPhenoAge^39^, PhenoAge PC1^40^, and DunedinPACE^41^. In addition, Elysium Health provided estimates from its proprietary Index clock and nine system-specific aging clocks. Index is a principal component-based epigenetic age predictor previously described^42^, while the system-specific clocks were previously applied by Norheim et al.^8^ and are based on the Systems Age framework described by Sehgal et al.^43^. Age-based clock estimates were expressed in years, whereas DunedinPACE estimates the pace of biological aging. DunedinPACE estimates were derived from incomplete coverage of the CpG sites included in the original algorithm. Previous validation demonstrated substantial agreement with the original predictor^42^.

For measures expressed in units of biological age, age acceleration was calculated separately for each measure. A linear regression model of biological age on chronological age was fitted using baseline measurements from all available participants. The resulting baseline model was subsequently applied to both baseline and post-intervention measurements. Age acceleration was defined as the difference between the observed biological age and the biological age predicted from chronological age. Positive values therefore indicated a biological age older than expected for chronological age, whereas negative values indicated a younger biological age. DunedinPACE was analyzed on its original scale because it estimates the pace of biological aging rather than biological age in years. For all measures, intervention-associated change was calculated as the post-intervention value minus the baseline value. Analyses were restricted to participants with paired baseline and post-intervention measurements.

Facial age (FaceClock) and mortality risk (FaceRisk) were estimated using previously developed deep-learning models, as described elsewhere^29^. Briefly, FaceClock estimates chronological age from facial photographs, whereas FaceRisk generates a facial mortality-risk score using a model trained directly on time-to-event mortality data. The pretrained models were applied to photographs from the present study without model retraining. FaceClock age acceleration was derived analogously to the other biological-age estimates. FaceRisk was analysed as a continuous score, with higher values indicating greater predicted mortality risk.

Blood-based age was estimated using Aging.AI 2.0 (BloodAge 2.0) and Aging.AI 3.0 (BloodAge 3.0), previously developed deep-learning models that predict chronological age from routine clinical biochemical and hematological measurements. The models were applied to blood measurements obtained at baseline and post-intervention, yielding age estimates expressed in years^28,44^.

To facilitate comparison between aging measures with different numerical scales, changes were divided by the baseline standard deviation of the corresponding measure. The resulting standardized changes therefore represent pre-to-post changes in units of the baseline standard deviation, with zero indicating no change.

Between-group intervention effects were evaluated using analysis of covariance models with the post-intervention value as the dependent variable and the corresponding baseline value, sex and intervention group as independent variables. Estimated marginal means were used to compare each active intervention group with the control group. Reported P values were not adjusted for multiple comparisons. Within-group changes displayed in the heatmap were evaluated using two-sided Wilcoxon signed-rank tests against zero.

PBMC cell-type proportions (CD4^+^ T cells, CD8^+^ T cells, B cells, NK cells and monocytes) were estimated from DNA methylation data using EpiDISH with robust partial correlations and the centDHSbloodDMC reference, using 250 available reference CpGs. Analyses included 57 participants with paired baseline and post-intervention samples. Between-group differences in cell-type proportions were assessed using ANCOVA adjusted for baseline values and sex. To assess whether changes in cell composition influenced DNAm aging estimates, the original ANCOVA models were repeated with additional adjustment for changes in CD8^+^ T-cell, B-cell, NK-cell and monocyte proportions, using CD4^+^ T cells as the reference category. All intervention effects were compared against CTRL.

### RNA extraction

PBMC pellets were thawed on wet ice, and total RNA was extracted using TRIzol reagent (Thermo Fisher Scientific, 15596026) according to the manufacturer’s protocol. Briefly, 1 ml TRIzol was added to each sample and mixed until fully dissolved, followed by the addition of 200 µl chloroform (Sigma-Aldrich, 1.02445). After vigorous shaking and incubation for 5 min at room temperature, samples were centrifuged at 12,000 × g for 15 min at 4 °C, and the aqueous phase was transferred to new tubes. RNA was precipitated with 500 µl isopropanol (Sigma-Aldrich, 109634) and washed with 70% ethanol. Pellets were resuspended in 450 µl absolute ethanol and 50 µl 3 M sodium acetate (Sigma-Aldrich, S7670) and stored at −80 °C overnight. The next day, samples were centrifuged at 21,000 × g for 15 min at 4 °C, washed once more with 70% ethanol, air-dried, and eluted in 30 µl nuclease-free water. RNA concentration and purity were assessed using a BioTek Epoch microplate spectrophotometer with Gen5 software (v1.10). RNA samples were stored at −80 °C until sequencing.

### RNA sequencing

RNA sequencing was performed by BGI Genomics (Shenzhen, China) on 105 PBMC samples. Strand-specific mRNA libraries were sequenced using 100-bp paired-end reads on the DNBSEQ platform, generating approximately 4 Gb of clean data per sample. Transcript abundance was quantified using Salmon against the GRCh38 reference. Following quality control and restriction to complete paired samples, the final analysis included 90 samples from 45 participants (CTRL, *n* = 8; EXE, *n* = 13; NR, *n* = 14; TRE, *n* = 10). Gene-level differential expression was analyzed using DESeq2. Genes were ranked by the DESeq2 Wald statistic for GSEA Preranked analysis against Gene Ontology Biological Process gene sets. Enriched pathways (FDR < 0.25) were visualized using Cytoscape (v3.10.0) with EnrichmentMap, AutoAnnotate, WordCloud and clusterMaker2.

### Protein quantification

Protein pellets were dissolved in 250 µl 5 M NaOH, heated at 80 °C for 30 min with intermittent vortexing and centrifuged at 12,000 × g for 10 min. Supernatants were diluted 1:200, and protein concentrations were measured in triplicate using the Pierce BCA Protein Assay Kit according to the manufacturer’s microplate protocol (Thermo Fisher Scientific, 23225) after incubation at 30 °C for 30 min.

### NAD^+^ quantification

Whole-blood NAD⁺ was quantified using a fluorometric enzymatic cycling assay adapted from Jensen et al.^45^. Whole blood (100 µl) was mixed with 1 ml 0.5 M perchloric acid and stored at −80 °C. After thawing on ice and centrifugation at 12,000 × g for 10 min, supernatants were diluted 1:200 in 100 mM sodium phosphate buffer, pH 8.0. Samples and β-NAD standards were analysed in triplicate following 1:1 addition of reaction mixture containing 100 mM sodium phosphate buffer (pH 8.0), 5 mM nicotinamide, 1% ethanol, 0.065 U/ml charcoal-treated diaphorase, 45 U/ml charcoal-treated alcohol dehydrogenase, 5 µM flavin mononucleotide and 1.25 µg/ml resazurin (final assay concentrations). Fluorescence was measured at 544/580 nm for 15–30 min, and NAD⁺ concentrations were determined from the linear range of the standard curve. NAD⁺ concentrations were normalized to protein content and expressed as pmol NAD⁺ per mg protein.

### Statistical methods

Sample size was determined a priori based on the prespecified primary outcome, plasma IL-6 at 12 weeks. The calculation compared the three active intervention groups pooled (EXE, NR and TRE) with CTRL. Based on prior studies reporting moderate-to-large effects of aerobic exercise, time-restricted eating and NR supplementation on circulating IL-6^7,10,46,47^ a large, standardized effect size (Cohen’s d = 0.80) was assumed. With a two-sided α = 0.05 and 80% power, 68 participants were required (51 pooled intervention and 17 control). To allow for an anticipated 15–20% attrition or treatment-discontinuation rate, the recruitment target was increased to 80 participants, with 20 participants assigned to each of the four study arms. No interim analyses were planned. Intervention effects were evaluated separately for EXE, NR and TRE relative to CTRL to preserve their distinct biological effects. The protocol-specified intention-to-treat mixed-model analysis was replaced by per-protocol ANCOVA to assess intervention responses among adherent participants.

Statistical analyses were performed in R (version 4.5.0). Continuous variables are summarized as mean (SD) or median (IQR), as appropriate. Participants were analyzed according to their intervention group. Within-group changes from baseline to post-intervention were assessed using paired t-tests; Wilcoxon signed-rank tests were used for the within-group biological-aging comparisons displayed in the standardized heatmap. Between-group intervention effects were evaluated primarily using ANCOVA, with the post-intervention value as the dependent variable, the corresponding baseline value as a continuous covariate, and sex and intervention group as categorical fixed effects. Estimated marginal means were used to compare each active intervention group (EXE, NR and TRE) with CTRL. For fat and muscle mass, ANCOVA models were fitted to log-transformed baseline and post-intervention values, with estimates presented as percentage differences. BMI and other continuously scaled outcomes were analyzed on their original scale unless otherwise specified.

For biological-aging measures, age acceleration was analyzed using ANCOVA with post-intervention age acceleration as the dependent variable and baseline age acceleration, sex and intervention group as independent variables. DunedinPACE was analyzed on its original scale. Changes displayed in the biological-aging heatmap were standardized by dividing the pre-to-post change by the baseline standard deviation of the corresponding measure. Individual intervention comparisons were not adjusted for multiple testing.

Associations between MVPA and intervention-related changes in outcomes within EXE were evaluated using linear regression, with effect estimates expressed per 100 min week⁻¹ of MVPA. FDR correction was applied across these exploratory tests using the Benjamini– Hochberg procedure. Cross-sectional associations between chronological age and baseline outcomes were evaluated using linear regression, including sex as a covariate in the adjusted analyses, with Benjamini–Hochberg correction across outcomes.

For the cross-domain analysis, Spearman correlations were calculated between pre-to-post changes in 85 outcomes, pooling participants across intervention groups and using available paired observations for each outcome pair. Across the 3,570 unique pairwise correlations, P values were adjusted using the Benjamini–Hochberg procedure. Associations with |ρ| ≥ 0.40 and FDR-adjusted P ≤ 0.10 were retained for visualization and descriptive analysis.

To examine exploratory associations between transcriptional and phenotypic changes, single-sample pathway activity was estimated using ssGSEA (GSEApy v1.1.13; rank normalization; gene-set size 5–2,000) with the 50 MSigDB Hallmark gene sets (h.all v2026.1). Changes in pathway activity from baseline to week 12 were compared with corresponding changes in 97 clinical, biochemical, physiological, functional and biological-aging outcomes among 45 participants with paired RNA-sequencing data (CTRL, n = 8; EXE, n = 13; NR, n = 14; TRE, n = 10). Age-based biological-aging measures were expressed as age acceleration, as described above. Associations were evaluated using rank-based partial correlations adjusted for intervention group, chronological age, sex and baseline outcome values. For each pathway–outcome pair, ranked pathway changes were modeled against ranked outcome changes and covariates. Partial correlations were calculated as the signed square root of the partial R², with P values derived from F tests. Benjamini–Hochberg correction was applied across 4,850 pathway–outcome comparisons. Overall transcriptomic–phenotypic coupling was assessed using the mean squared partial correlation across all pairs, compared with a null distribution generated from 2,000 permutations of participant pathway profiles within intervention groups. Hallmark gene sets were additionally grouped into ten clusters according to correlations in their adjusted changes using average-linkage hierarchical clustering, independently of phenotypic outcomes. Outcomes were classified into 15 measurement domains. Associations between the resulting 150 pathway-cluster x outcome-domain combinations were evaluated using mean squared partial correlations and 20,000 within-group permutations, with Benjamini–Hochberg correction across combinations. An FDR threshold of q < 0.25 was used to identify exploratory cluster-level associations. Differences in pathway–outcome correlations between intervention groups were assessed using Cochran’s Q statistics on Fisher z-transformed within-group correlations, with overall heterogeneity evaluated using 2,000 permutations of intervention labels.

## Data availability

Requests for data supporting the findings reported in this study should be directed to the corresponding author and will be reviewed individually, subject to institutional approval and applicable data-protection requirements. Individual participant-level clinical data and DNA methylation data cannot be shared publicly owing to participant confidentiality and restrictions under informed consent. For RNA sequencing data, DESeq2-normalized gene-level counts from individual PBMC samples are available at https://doi.org/10.5061/dryad.g4f4qrg6v.

## Code availability

R scripts used for statistical analyses and data visualization are available from the corresponding author upon request.

## Acknowledgments

We thank all study participants for their time and commitment.

## Contributions

K.T.N. and M.S.-K. conceived and designed the study. M.B. coordinated the trial, collected participant data, performed laboratory experiments and statistical analyses, and drafted the manuscript. H.A. and S.R. contributed to participant recruitment and data collection. S.R. and J.B.G. performed laboratory experiments. B.O. and M.B. conducted the NAD⁺ analyses, with B.O. additionally supervising laboratory work. M.P. performed the gait analyses. A.D. and M.B.E. performed the RNA-sequencing analyses. I.H. performed the facial-aging analyses. G.M. supervised laboratory work. M.S.-K. supervised the study. All authors reviewed and approved the final manuscript.

