## Supplementary figures for "Comparative effects of exercise, nicotinamide riboside and time-restricted eating on aging-related biology in older adults: a randomized, controlled trial"

Present addresses:
[†]Department of Clinical Research, Copenhagen University Hospital Amager & Hvidovre, 2650, Hvidovre, Denmark.
[#]School of Biotechnology and Biomolecular Sciences, University of New South Wales, Kensington, NSW 2052, Australia.
[$]Tracked Biotechnologies LLC, Manassas, VA, USA, and Tracked Biotechnologies ApS, Copenhagen, Denmark.
[§]Senescope, Santa Cruz, CA 95060, USA.

**Contains:**
Supplementary figures 1,2 and 3

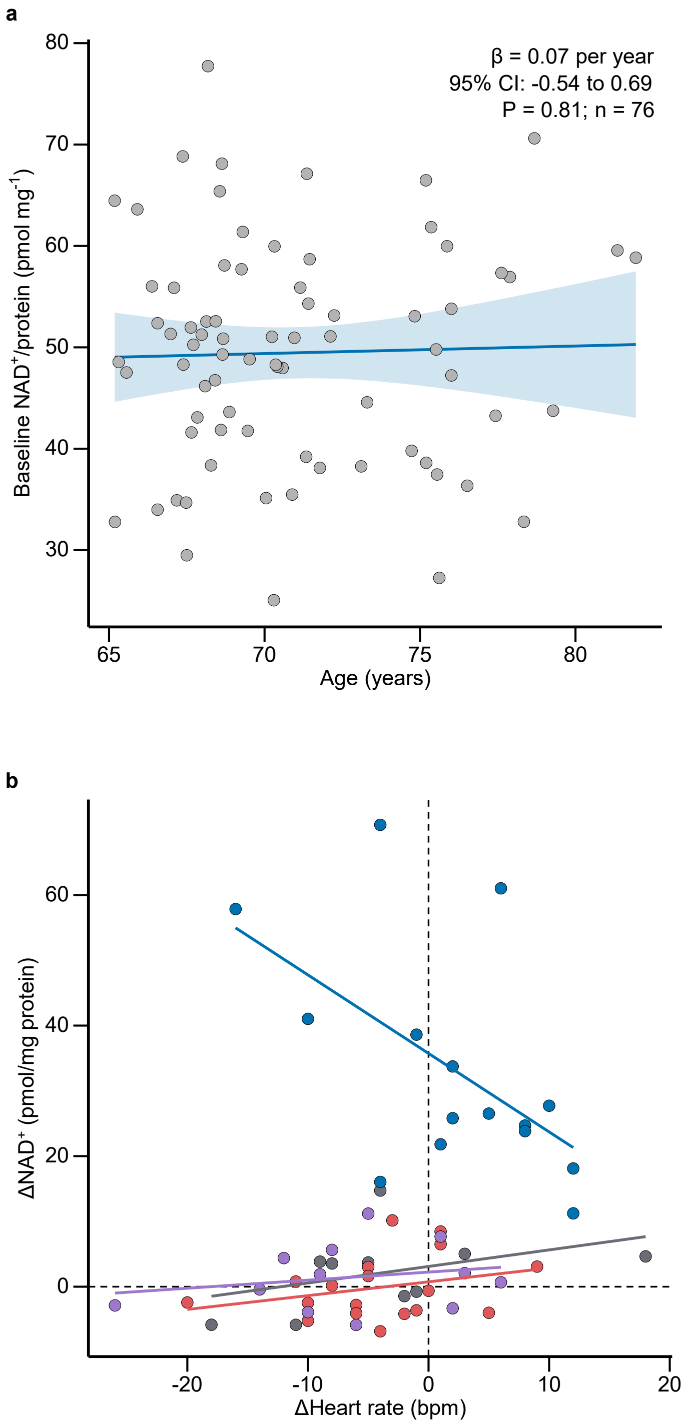


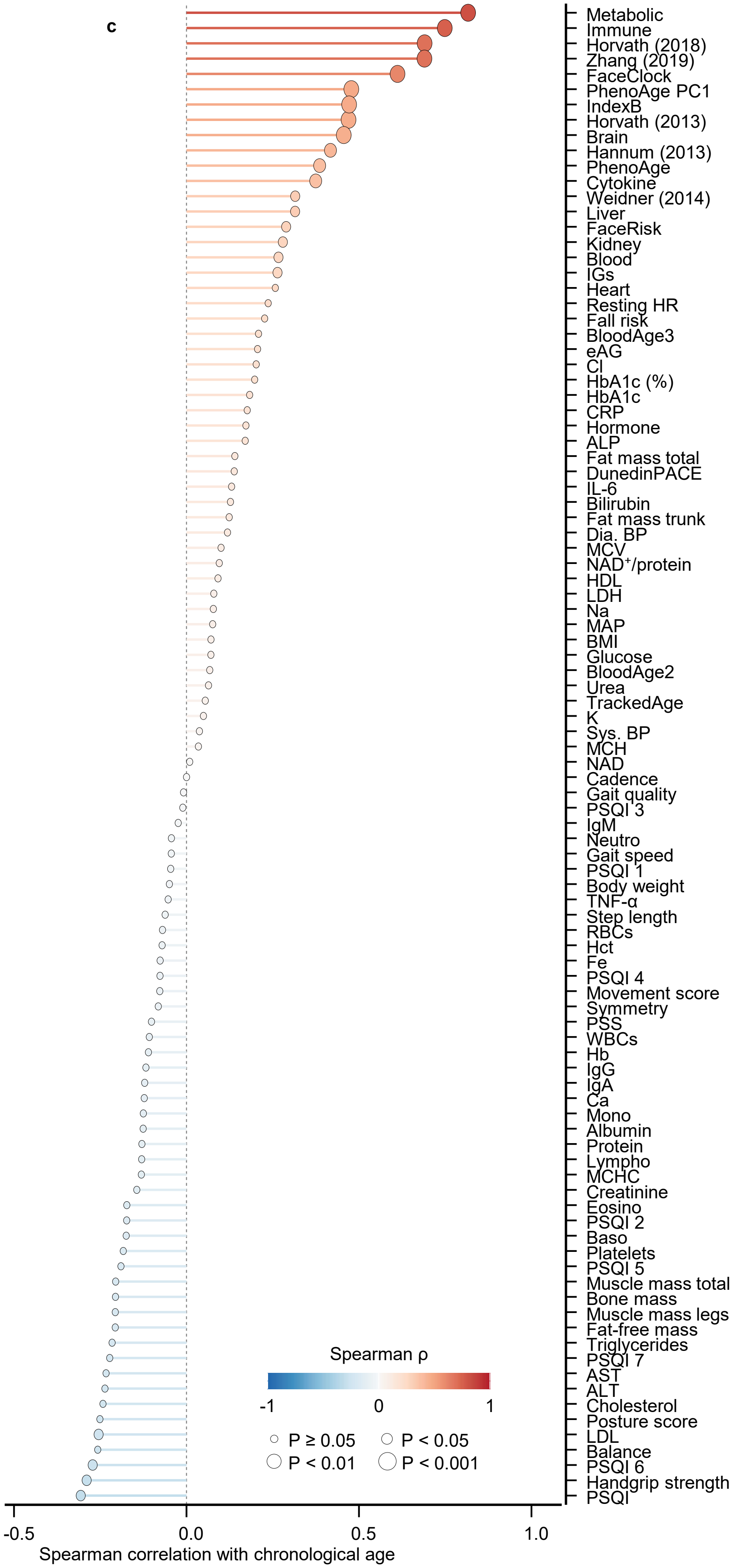


**Supplementary Figure 1 | Associations between NAD⁺, resting heart rate, chronological age and study outcomes.**
**a**, Association between baseline NAD⁺ and chronological age (n = 76). Solid line shows linear regression with 95% CI. **b**, Associations between pre- to post-intervention changes in resting heart rate and whole-blood NAD⁺, with intervention-specific linear regression fits (CTRL, grey; EXE, red; NR, blue; TRE, purple). Dashed lines indicate no change. **c**, Spearman correlations between chronological age and baseline study outcomes. Point color indicates correlation direction and magnitude (blue, negative; red, positive), and size indicates the unadjusted two-sided P value.

**a**

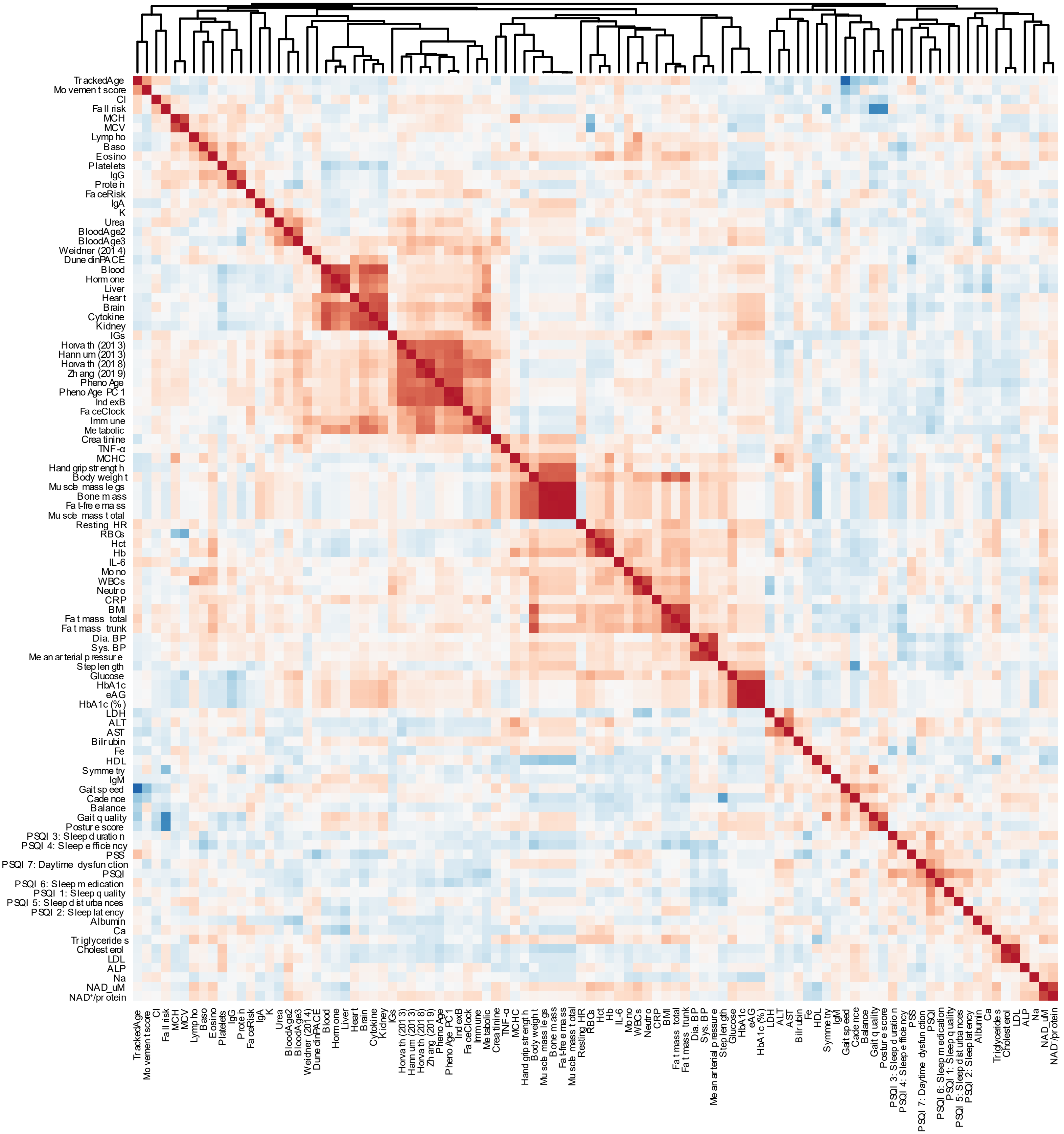


**Supplementary Figure 2 | Spearman correlation matrices of study outcomes at baseline and their changes following intervention.**
**a**, Pairwise Spearman correlations between baseline study outcomes. Correlations were calculated using pairwise complete observations. Outcomes were hierarchically clustered using average linkage based on 1 − Spearman ρ. Colors indicate correlation direction and magnitude (blue, negative; red, positive).

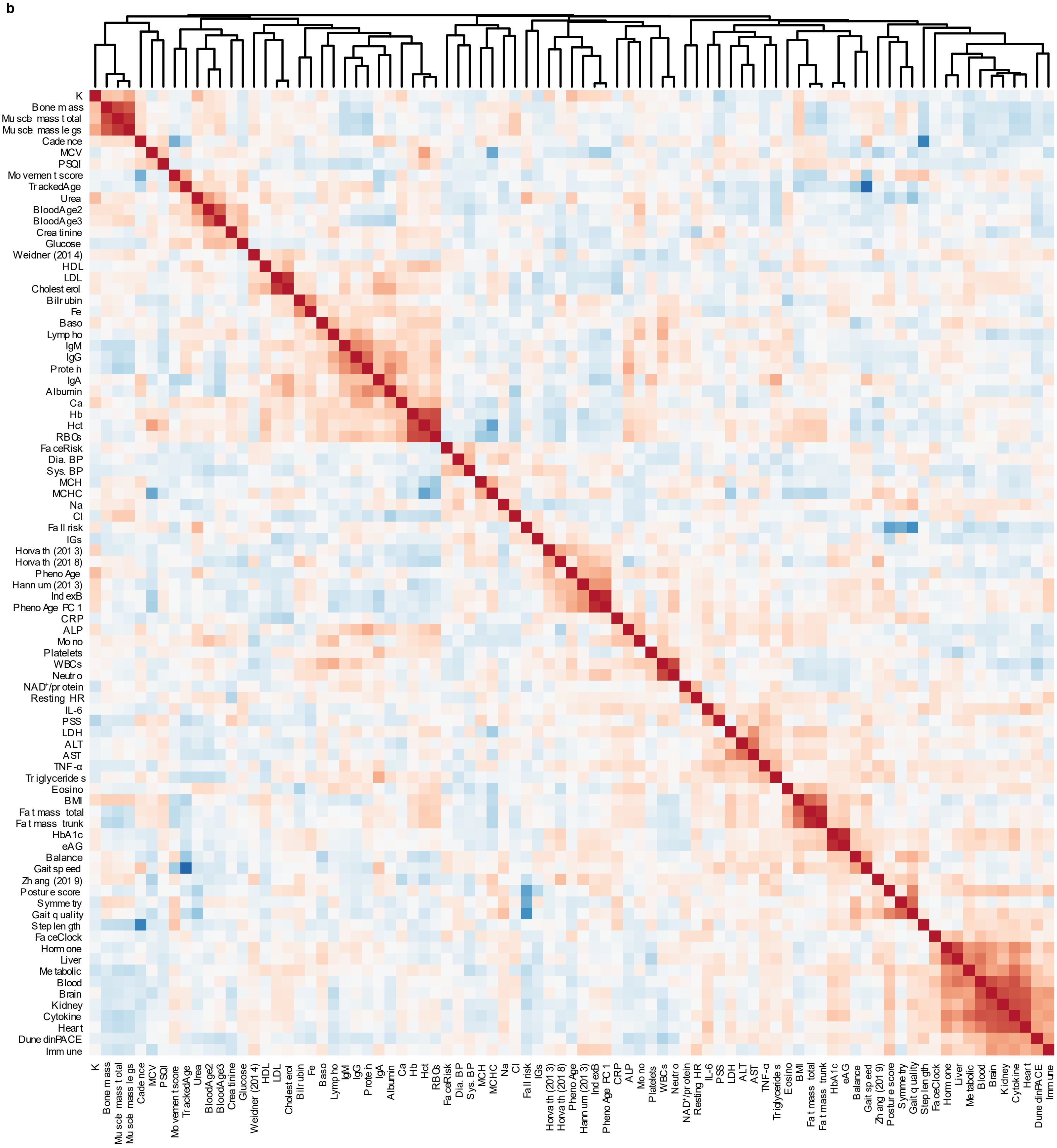


**Supplementary Figure 3 | Spearman correlation matrices of study outcomes at baseline and their changes following intervention.**
**a**, Pairwise Spearman correlations between pre- to post-intervention changes, pooled across intervention groups. Correlations were calculated using pairwise complete observations. Outcomes were hierarchically clustered using average linkage based on 1 − Spearman ρ. Colors indicate correlation direction and magnitude (blue, negative; red, positive).
